# Enterovirus D68 in Europe, 2025: continent-wide predominance of subclade A2/D and shift from pediatric to adult cases

**DOI:** 10.64898/2026.10.23.26366309

**Authors:** Robert Dyrdak, Georg Navér, Nadia Neuner-Jehle, Alejandra Gonzalez Sanchez, Yemisi Adedeji, Lili Andersson-Li, Cristina Andrés, Andrés Antón, Brynja Armannsdottir, Kafayat Arowolo, Aishwarya Arunagirinathan, Guðrun Erna Baldvinsdottir, Natasa Berginc, Kelly Bicknell, Maxime Bisseux, Sandra Broddesson, Concetta Castilletti, Pierluigi Congedo, Payton Crossley, Lize Cuypers, Natascha de Graaf, Anna Eis-Hübinger, Rebecca Feletti, Licínia Gomes, Raquel Guiomar, Marion Jeannoel, Georgina McAllister, C. Patrick McClure, Adam Meijer, Sofie Midgley, Audrey Mirand, Richard Molenkamp, Carmen Muñoz-Almagro, Jean-Luc Murk, Davide Oggioni, Massimo Oggioni, Elena Pariani, Laura Pellegrinelli, Amaresh Perez-Arguello, Miroslav Petrovec, Antonio Piralla, Antonino Maria Guglielmo Pitrolo, Elena Pomari, Morten Rasmussen, Marijke Reynders, Lieuwe Roorda, Isabelle Schuffenecker, Leo Smeets, Alison Tedcastle, Kate Templeton, Tina Uršič, Jaco J. Verweij, Jorgina Vila, Karin von Eije, Elke Wollants, Joakim Øverbø, Javier Martin, Kim Benschop, Heli Harvala, Thea Kølsen Fischer, Emma B. Hodcroft

## Abstract

**Background:** Enterovirus D68 (EV-D68) has predominantly caused infections in children, and detected cases have mainly been infected by subclade B3. In 2025, however, a substantial number of infections in adults was noted.

**Aim:** To detail the molecular epidemiology of the 2025 season in Europe.

**Methods:** Case and sequencing data was collected from members in the European Non-Polio Enterovirus Network (ENPEN). The participating sites included both clinical and public health laboratories.

**Results:** Data was collected from 13 countries at 23 sites. There were 187 clinical cases infected with EV-D68 and 127 detections in wastewater. Age data were available for 76% (143/187), among which 90 cases were adults. VP1-based genotyping showed subclade A2/D to predominate in all age groups and across all participating countries, as well as in the sewage sampling performed at one site. Two interclade recombinant sequences were detected among 22 whole genome sequences.

**Conclusion:** The observed shift towards adult cases and subclade A2/D predominance warrants continued monitoring to determine whether this represents a sustained epidemiological change. The detection of interclade recombinant variants highlights a need to supplement regular VP1-based genotyping with whole-genome sequencing.

## Introduction

In late 2025, communication in the European Non-Polio Enterovirus Network (ENPEN) indicated an unusual epidemiological pattern of enterovirus D68 (EV-D68) circulation with a substantial number of cases in adults and general predominance of subclade A2/D.

EV-D68 is classified in clades A through C based on phylogenetic analysis of the VP1 gene. Subclade A2 is further subdivided by some authors into subclades D1 and D2 [1]. Previous outbreaks have mostly involved children, with clade B being the most common, whereas when detected in adults, EV-D68 mainly belongs to clade A2 [1, 2, 3, 4, 5, 6].

Molecular surveillance of EV-D68 circulation in the 2024 season indicated subclade B3 predominating with subclade A2/D co-circulating at lower levels in many regions, including North America [7] and Asia [8]. In contrast, in Europe an increase in EV-D68 cases was observed in outbreaks of acute respiratory infections in northern Italy [9, 10], Spain [11, 12] and France [13], where sequence data show subclade A2/D predominating with B3 co-circulating at lower levels. Notably, these European outbreaks were also associated with an increase in adult cases. Also in China a study on pediatric patients noted an upsurge of subclade A2/D in 2024 [14]. Furthermore, data from season 2025 in the northeast US suggest an exclusive A2/D circulation [15]. Here, we detail the molecular epidemiology of EV-D68 in Europe, 2025.

## Methods

Data were collected from 13 European countries in 23 sites participating in the European Non-Polio Enterovirus Network (ENPEN). Hereinafter, the sites are designated by the two-letter ISO country-code and sequential number, see supplement table for details. Of the participating sites, 22 sites analyzed clinical samples and one site (UK3) performed environmental wastewater sampling. Of the participating sites, 15 were hospital-based (BE1, BE2, DE1, ES1, ES2, FR1, FR2, IT2, IT3, IT4, NL2, SE1, SI2, UK1, UK2), 7 were public health institutes (DK1, IT1, NL1, NO1, PT1, SI1, UK3), and one site had both tasks (IS1). One of the hospital-based laboratories was dedicated to pediatric patients (ES1). Surveillance approaches included genotyping of specimens from clinical routine EV/RV diagnostics at 11 sites (BE2, ES1, ES2, FR1, FR2, IS1, IT2, NL2, SE1, SI2, UK1), from ILI/ARI surveillance at 7 sites (DE1, IT1, IT3, IT4, PT1, SI1, UK2), from both these sources at 2 sites (BE1, DK1), at one site from a dedicated enterovirus surveillance (NL1a) and an ILI/ARI general practice sentinel surveillance (NL1b), and at one site from enterovirus and acute flaccid paralysis surveillance along with collection of positive results for EV-D68-specific PCR from clinical laboratories (NO1). See S1 for details.

The primary method of EV-D68 detection was specific PCR at 6 sites (BE1, FR1, PT1, SI2, UK1, SE1), enterovirus sequencing at 5 sites (ES1, ES2, FR2, NL1, UK2), and a combination of these methods at 11 sites (BE2, DE1, DK1, IS1, IT1, IT2, IT3, IT4, NL1, NL2, NO1, SI1). Across the sites, there were 4 EV-D68 specific PCR assays used, based on the published protocols by Piralla et al. [16] (BE2, DK1, IT2, IT3, SE1, SI1), Poelman et al. [17] (BE1, DE1, FR1, IT1, IT4, NL1, NL2, PT1, UK1), Bragstad et al. [18] (IS1), and Wylie et al. [19] (SI2). For PCR detection, one site (NO1) reported positive result notifications from hospital-based laboratories without information about the assays used there or denominator data for number of samples analyzed.

Testing for EV-D68 was done according to local routines and testing algorithm varied across sites. EV-D68 testing could thus be performed e.g. on all samples tested for enterovirus/rhinovirus (BE1, BE2), on all respiratory samples during autumn season (FR1), or on a random subset of samples positive for enterovirus/rhinovirus during autumn season (SE1).

Wastewater samples were collected from 13 locations across the UK and analyzed directly from sewage concentrates using established protocols incorporating both generic and clade-specific EV-D68 PCR assays, followed by Oxford Nanopore Technologies (ONT) sequencing [20]. Reads generated from the generic EV-D68 PCR assay were assigned to clades B3 or A2/D to estimate their relative proportions within each sample.

Of the 17 sites that submitted sequences, 9 sites submitted partial VP1 sequences (BE1, DK1, ES2, IS1, IT1, IT4, NL2, PT1, SI2), 3 sites submitted complete VP1 sequences (DE1, UK2, UK3), 3 sites submitted whole genome sequences (FR2, IT2, SE1), and 2 sites (BE2, NL1) submitted both partial VP1 and whole genome sequences. Genotyping was done using the EV-D68 dataset in Nextclade (v3.21.2) [21].

For the analysis of age and clade over time and as context for the phylogenetic analysis, publicly available VP1 sequences downloaded from BV-BRC 10 May 2026 (*≥* 700bp until 2023, *≥*300bp for sequences from 2024 and 2025) were included alongside the data generated here, with ages provided alongside the sequences or taken from the literature as previously described [2]. To look at overall European clade trends by year, all VP1 sequences *≥*300bp between 2013-2025 were included. For phylogenetic analysis, sequences were analyzed with the Nextstrain pipeline as previously described [2]. Initial assessment for potential recombination was done on whole-genome sequences in Nextclade [22], and full recombinant analysis on selected sequences was done using RDP5 [23].

Statistical comparisons of age distributions across years were performed using Fisher’s exact test, only including sequences where age could be classified as *≥*18 or <18 years. Further methodological details are provided in Supplementary Methods. Code to plot pie charts of wastewater detection (Fig. 1), figures related to age analysis (Fig. 4, Fig. 2, Fig. S4) and modify the phylogenetic tree to highlight the sequences generated in the study (Fig. 3) was generated with assistance from Claude Code Haiku 5.5.

**Fig 1.**
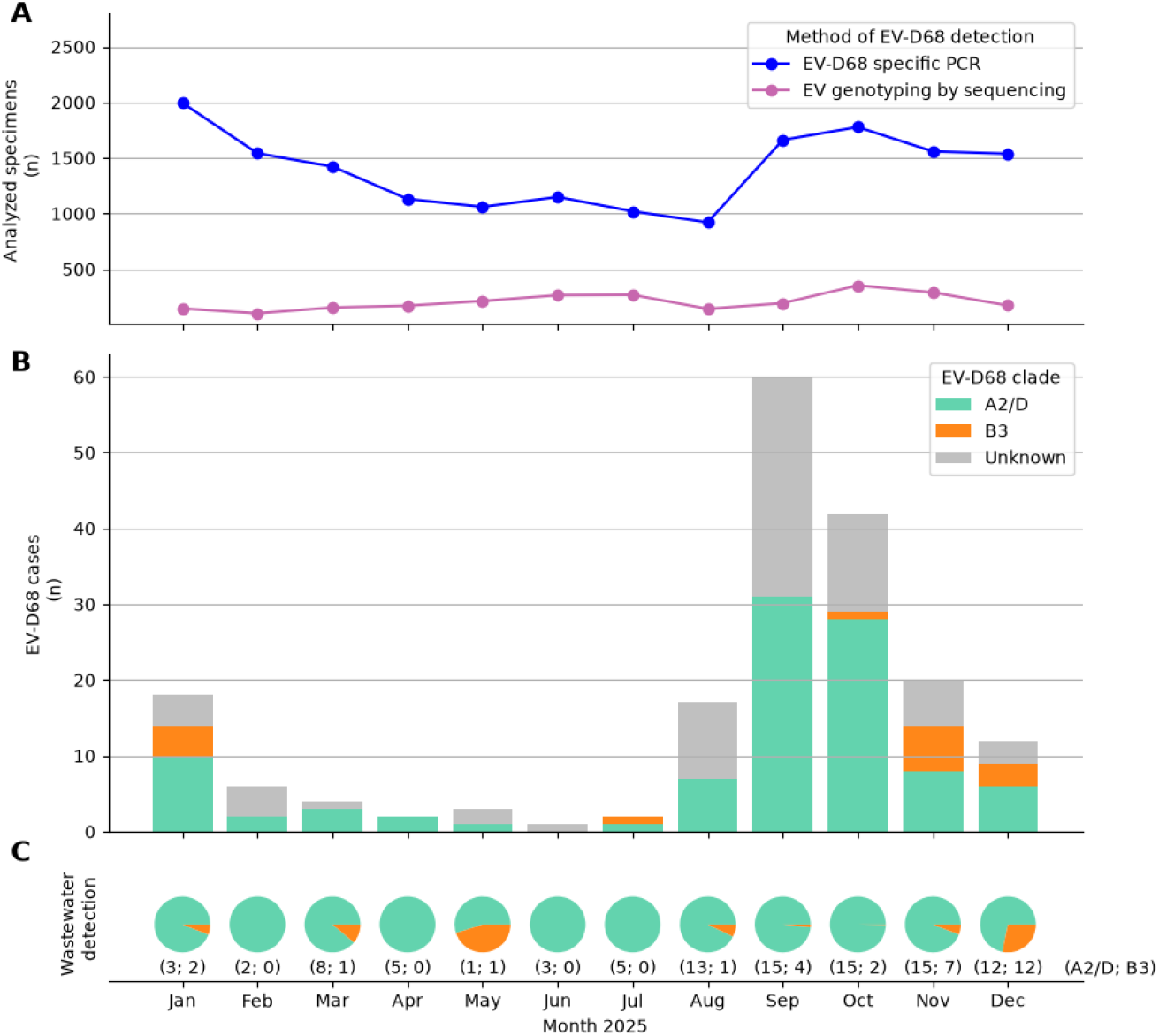
Testing and results for EV-D68 per month 2025. A: Number of respiratory specimens across all sites analysed by EV-D68 specific PCR or using enterovirus genotyping by sequencing. B: Number of cases with detected EV-D68 across all sites coloured by subclade. C: Detection of EV-D68 in wastewater samples in the UK. Pie charts show the monthly mean proportions of Oxford Nanopore Technologies (ONT) sequence reads assigned to clades B3 and A2/D following generic EV-D68 PCR, calculated by giving each sample equal weight. Numbers below the pies indicate the number of distinct strains detected for each clade that month (A2/D; B3).

**Fig 2.**
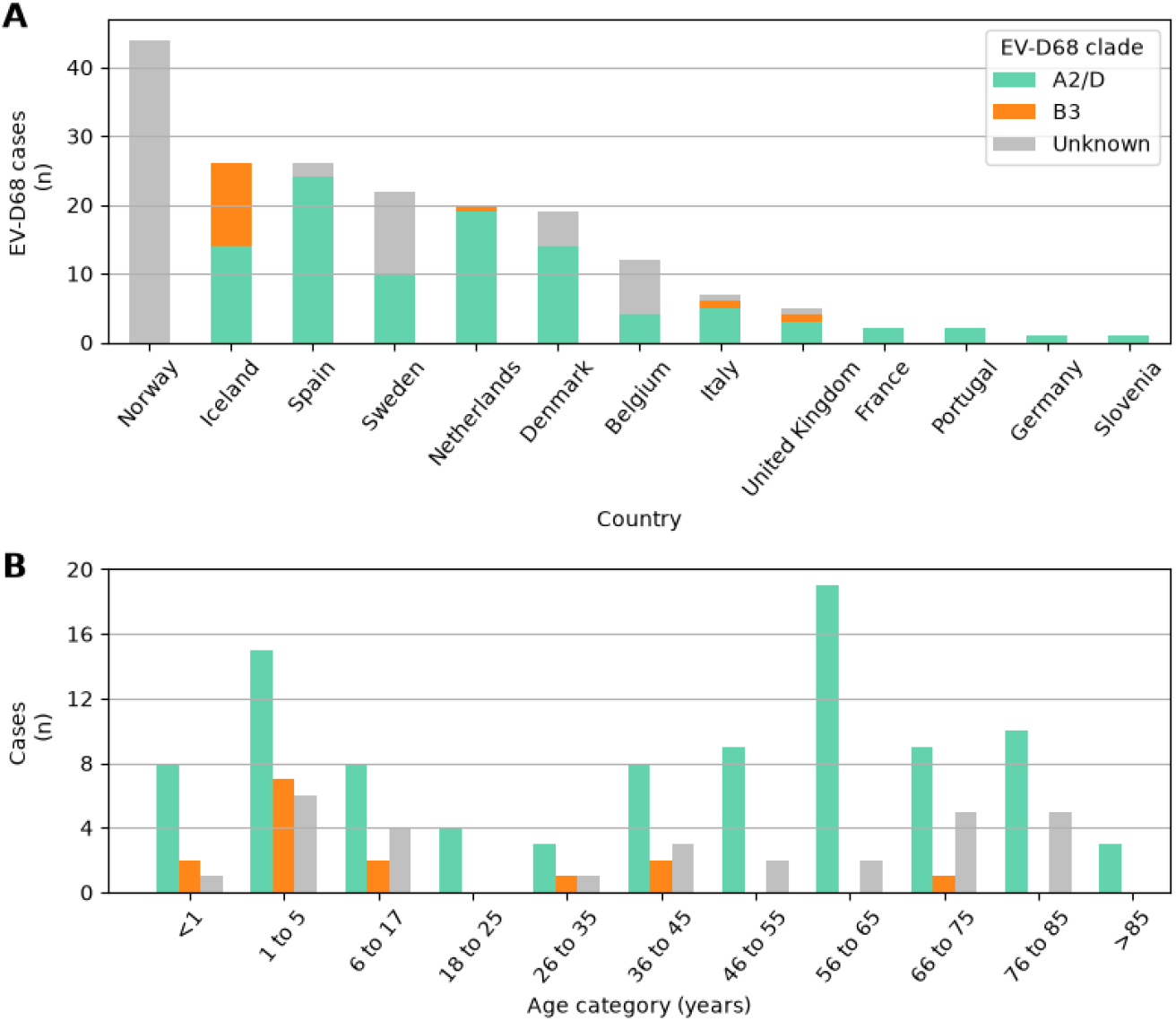
EV-D68 Cases by Age Group. A: Number of detected cases per country colored by genotype. B: Number of detected cases per age category. There were 44 cases that did not have age information.

**Fig 3.**
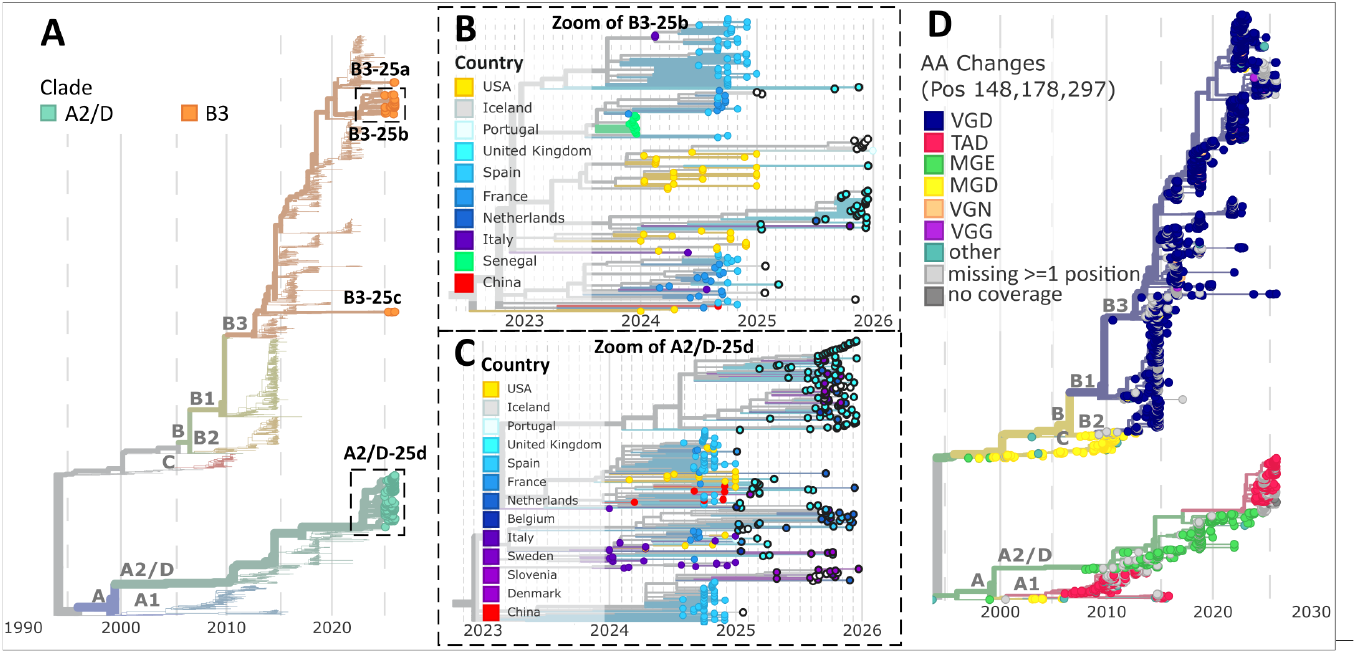
European 2025 EV-D68 Sequences in Global Phylogeny. **A:** Overview of the 2025 European EV-D68 sequences within a phylogeny comprising all publicly available sequences, identifying three major clusters to which the European sequences belong. **B & C:** Enlarged views of cluster B3-25b and A2/D-25d, respectively, with sequences generated in this study outlined in thick black. **D:** Phylogeny annotated with amino acid substitutions at VP1 positions 148, 178, and 297 This analysis demonstrates that the recently emerged A2/D lineage circulating since 2024 shares the same amino acid substitutions at these positions as the A1 subgenogroup (highlighted in red), which has not been detected since 2015

**Fig 4.**
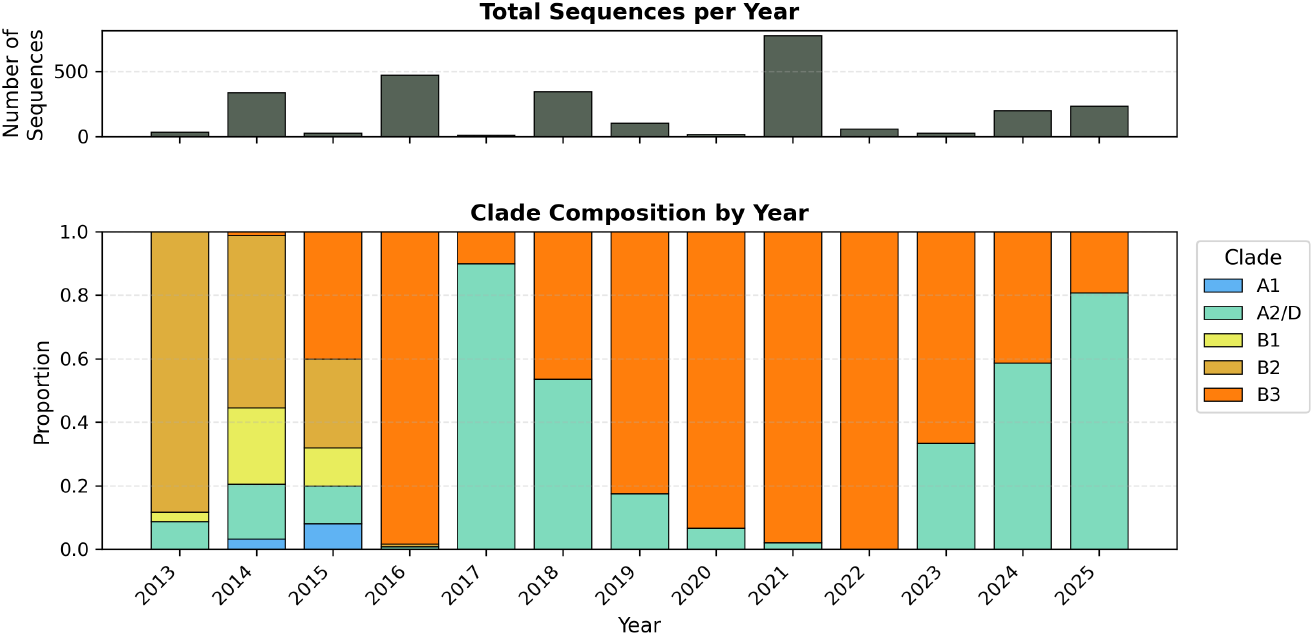
Sequence count and subclade of VP1 sequences from Europe 2013-2025. European sequences with at least 300bp of VP1, grouped by subclade and year. Subclade B (B1, B3) have historically predominated in each season, with marked exceptions in 2018 and since 2024 to present.

## Results

In total, 187 human infections with EV-D68 were detected, alongside 127 EV-D68 detections in wastewater samples. cases of EV-D68 infection were identified, and 127 detections in wastewater samples. With the exception of one site that provided data only on positive PCR results, the denominators for EV-D68 PCR and NPEV sequencing on EV/RV-positive samples were 16,777 and 2,486 respiratory specimens, respectively (Fig 1A). Of these, the vast majority of EV-D68 specific PCR tests was performed in Belgium (n=13,086), followed by the Netherlands (n=1,646) and France (n=1,072), whereas most NPEV sequencing was done in France (n=888), the Netherlands (n=624), and Spain (n=522).

More than half (n=102) of the EV-D68 positive cases were detected in September and October (Fig 1B). The peak of detected cases during the late summer and autumn period in 2025 was mirrored by a similar increase in the EV-D68 detection in wastewater at the UK site performing environmental testing (Fig 1C). In August through December, EV-D68 was detected in wastewater at 11 to 13 locations in the UK, up from 1 to 7 locations in January through July.

Sequence data were available for 61% (114/187) of EV-D68 positive cases, of which 22 sequences were whole genome, 7 were complete VP1 sequences, and 85 were partial VP1 sequences. Among cases with sequence data, based on the VP1 segment EV-D68 belonged to subclade A2/D in 87% (99/114) and to subclade B3 in 13% (n=15), and subclade A2/D predominated among sequences across the countries (Fig 1A).

Age data was available for 76% (143/187) of EV-D68 positive cases, and for all cases with sequence data available. There was a bimodal age distribution, with peaks in the age groups 1 to 5 years (20% (28/143)), and 56 to 65 years (15% (21/143)) (Fig 2B). Furthermore, clade A2/D predominated in all groups and was the only clade found in the age groups of 46 years and above, with the exception of 1 case from clade B3.

Likewise, the wastewater surveillance in UK showed an increase in the prevalence of EV-D68 detections in wastewater during the period of peak clinical case detection, with a clear predominance of subclade A2/D. Genotyping was performed in 12 of 13 countries, and clade A2/D was detected in all 12, with co-circulation of clade B3 was present in 4 countries (Iceland, Italy, the Netherlands, and United Kingdom).

### Phylogenetic analysis

From a starting dataset of 241 VP1 sequences (114 clinical and 127 wastewater), 11 were excluded from analysis due to length (textless 300bp), resulting in a final set of 230 sequences from this study included in the phylogenetic analysis (103 clinical and 127 wastewater). Due to extremely high mutation density, some sequences had up to 75 bases trimmed from the ends as suspected sequencing/primer noise (see Supp Methods)

The 2025 European sequences were mostly closely linked to samples from the previous year, both from Europe and the USA, with a few links to China. The interactive Nextstrain phylogeny can be viewed at https://nextstrain.org/community/enterovirus-phylo/evd68-2025/vp1.

The 230 sequences in the study clustered in 4 distinct areas of the tree (Fig. 3 A): 2 from a B3 subgenogroup cluster that circulated primarily in 2022 (B3-25a), 40 within a larger cluster of the B3 subgenogroup expanding from more recent sequences (B3-25b), 3 sequences within a small cluster of the B3 subgenogroup (B3-25c), and 185 sequences within the A2/D subgenogroup (A2/D-25d).

Subgenogroup B3-25a connects to sequences from 2024 in the US and Spain. Among B3-25b, most sequences (25/40) link to 2024 US circulation, others (5/40) derive from a 2024 France/Spain cluster, and the remainder (10/40) trace back to 2023 with no close connection to earlier samples. Most B3-25b sequences have only synonymous mutations, i.e. have no amino acid mutations. B3-25c clusters distantly from a 2023 China sample, with an MRCA dating to 2019. Among A2/D-25d sequences, many interleave closely with 2024 sequences from Spain, Italy, France and the US, with the majority (112/185) forming a distinct monophyletic subcluster which branched from the other samples in 2023. This subcluster has no defining amino acid mutations.

Overall among subclade B3, detections in wastewater constituted both of the sequences in the (B3-25a) cluster, 25 of 40 sequences in the B3-25b cluster, all three sequences in the B3-25c cluster. Among A2/D detections, 97 of the 185 samples were from wastewater. Wastewater samples were generally well-distributed throughout the phylogeny alongside clinical samples. However, wastewater constituted the entirety of the B3-25a and B3-25c clusters, which were phylogenetically distant from previous samples.

The branch leading to cluster A2/D-25d has four VP1 AA changes: M148T, G178A, A277T and E297D, with the inferred most common recent ancestor (MRCA) in late 2022.

Interestingly, of the four VP1 amino-acid changes present in the A2/D lineage circulating since 2024 (M148T, G178A, A277T and E297D), three of these (148, 178, 297) were also present in the A1 subgenogroup (Fig. 3, D), which was last detected in 2015.

Two of these positions, 148 and 297, are part of the surface-exposed DE-loop and C-terminus epitope, respectively, giving the A2/D-25d cluster slightly different DE-loop and C-terminus epitope patterns than previously circulating A2/D sequences.

Of the 22 whole genome sequences, two sequences (SWE/EV-D68_008/2025 and EV-D68/Netherlands/GP01/2025, accession numbers PZ672967 and PZ797495, respectively) were classified as subclade A2/D based on the VP1 segment but showed very long branches and were therefore investigated further. The analysis indicated both sequences were interclade recombinant variants, with both having the structural genes, thus including the VP1 gene, derived from subclade A2/D, whereas most of the non-structural genes were derived from subclade B3. The Swedish recombinant sequence (SWE/EV-D68_08/2025) had the breakpoint at approximately 3750bp, i.e. at the 2A/2B junction, whereas the breakpoint in the recombinant from the Netherlands was estimated to be near the N-terminus of the 2C gene, at approximately 4090bp. See supplement Fig S2 and Table S3 for details.

### Age trends

Historically during EV-D68 outbreaks in Europe since 2013, sequences from the B clade (particularly subclades B2 and B3) have dominated (Fig. 4). An interesting exception is in 2018, when A2/D and B3 sequences were at a similar proportion. Notably, since the winter of 2024, subclade A2/D sequences have been present at a higher level than B3 sequences. This increase in proportion of A2/D sequences appears to be correlated with an increase in adult cases (where ages are available) (see Supp Fig. S4).

The proportion of adults among EV-D68 cases with sequences and ages in this study (63.1%, 72/114) was significantly higher than in 2021 (12.0%, 25/208), the most recent B3-dominant outbreak year (Fisher’s exact test, p<0.001; OR=12.4, 95% CI: 6.9 to 23.0). Additionally, the proportion of adults was even significantly higher compared to 2018 (38.4%, 43/112), a previous A2/D-dominant year (p<0.001; OR=2.7, 95% CI: 1.5 to 4.9). These findings indicate that years characterized by predominance of subclade A2/D are consistently associated with a higher proportion of adult EV-D68 cases compared with B3-dominant years.

## Discussion

In this multicentre collaboration, we found that adults accounted for the majority of EV-D68 cases detected during the 2025 circulation season, and that subclade A2/D predominated across Europe and was identified in all age groups. This pattern is consistent with reports from northern Italy [9, 10], France [13], and Spain [11, 12] during the 2024 season, which also documented an increased proportion of adult cases alongside predominance of subclade A2/D. Since 2016, the EV-D68 circulation in Europe has been characterized predominantly by subclade B3 and a predominance of cases among children, with the notable exception of 2018, when increased circulation of subclade A2/D coincided with a higher proportion of adult cases. However, the proportion of adult cases observed in 2025 exceeded that reported in 2018. These findings raise the question of whether the apparent association between subclade A2/D predominance and increased detection of cases among adults, including older adults, will persist in future EV-D68 circulation seasons. Notably, including 2018, this is the second time that subclade A2/D dominance has been observed, raising the possibility that such upsurges may recur periodically rather than representing an isolated event.

A possible reason for the upsurge of subclade A2/D could be due to an escape of the virus from existing population immunity. The reappearance of VP1 mutations previously seen in the A1 subclade (2006-2015, Fig. 3 D) suggests these mutations may confer immune escape from the dominant B3 immunity that has accumulated since 2016. In those who previously encountered subclade A1, immunity may have waned; in those born after 2015, it is absent entirely, which could create conditions for a virus carrying these mutations to potentially gain fitness advantage. This is however speculative as data on population immunity against subclade A2 is missing. Although a serosurvey in the Netherlands showed a high level of population immunity against EV-D68 in both children and adults in sera from 2006–07 and 2015–16, with only minimal differences in titers to the prototypic Fermon strain compared to subclade B3, in that study titers against clade A were not measured [24]. Notably, the inferred most recent common ancestor of the current European subclade A2/D cluster was late 2022, thus indicating that these variants have circulated several years prior to the upsurge in 2024 and 2025. Thus, while it remains unclear why such an upsurge in A2/D was observed only in 2024-5, the relatively low circulation of A2/D between the 2018 upsurge and 2024 may have allowed population immunity to wane without being re-boosted by new exposures.

It is worth noting that in 2025 subclade A2/D predominated not only among adults but also in young children (Fig. 2 B), who should be largely immunologically naive. This suggests that the circulating subclade may be determined by population-level transmission dynamics rather than age-specific susceptibility patterns alone. While adults comprised over half of detected cases in 2025, and subclade A2/D was detected in 60 of 61 adults *≥*46 years in whom genotyping was performed, subclade A2/D also predominated among young children, who are likely to be immunologically naive. This can suggest that the circulation may be driven by population-level transmission dynamics, such that adult populations may have played a substantial role in seeding pediatric infections, as has been hypothesized previously [2]. Further work should focus on understanding these inter-generational transmission dynamics better, as they could have implications for surveillance strategies. Similarly, more serological data is needed on immunity against A2/D strains: a recent systematic review of age-stratified seroprevalence studies found only two datasets assessing antibodies against clade A or A2/D, both prior to the dominance of A2/D in 2018 [25].

Surveillance-based studies are always prone to biases in testing. However, the relatively sudden emergence of elevated adult proportions in 2018 and 2024-2025, rather than gradual increase, argues against testing practice changes as a cause for the observed correlation. More importantly, the consistent association between A2/D and adult case frequency in both 2018 and 2024-2025, separated by B3 dominance with low adult cases, suggests a clade-specific effect rather than surveillance artifact. Testing practices and awareness of adult EV-D68 infection were substantially lower in 2018 than 2024-2025, yet A2/D dominance and the association with adult cases was equally evident. The predominance of both A2/D and the relative increase in adult cases was additionally consistent across the participating countries and different surveillance strategies.

As surveillance based on clinical cases carries the risk of being biased towards symptomatic cases, environmental surveillance of wastewater allows for a more unbiased sampling of the circulation. Thus, the predominance of subclade A2/D also in the wastewater detections during the peak circulation supports the notion of an actual predominance of subclade A2/D rather than a sampling bias. However, the detections of subclade B3 in wastewater samples also indicate a significant unsampled diversity that could allow for novel clusters to emerge among cases in coming seasons. While wastewater samples were generally well-distributed throughout the phylogeny alongside clinical samples, the B3-25a and B3-25c clusters were composed entirely of wastewater sequences and were phylogenetically distant from the main circulating strains. This finding highlights the importance of wastewater surveillance to detect low-frequency variants that escape clinical detection.

We identified a recombinant sequence from Sweden that had an identical breakpoint to the five A2/D-B3 recombinant sequences identified in autumn 2025 in the US [15], indicating that this recombinant variant has circulated internationally. A second recombinant sequence from the Netherlands was also detected, however, this sequence had a different breakpoint, indicating a separate origin. Although the ability to detect the recombinant variants in this study was limited, as 81% (92/114) of clinical sequences were limited to the VP1 gene, recombinant EV-D68 variants do not seem to be the likely cause of the upsurge of clade A2/D as they were detected in only 2 of the 22 whole-genome sequences. Nevertheless, a novel combination of structural and non-structural genes might generate a variant with increased fitness, as exemplified by recombination being a founding event for most EV-A71 subgenogroups [26], and having been observed to correlate with outbreaks in echovirus types 9, 11, and 30 [27]).

Further studies would be needed to assess the biological properties of the EV-D68 recombinant variants. Still, the detections of these recombinant variants underscore the importance of complementing surveillance based on VP1 sequencing with whole-genome sequencing to allow such variants to be identified. In particular, recombinant generation becomes more likely when multiple lineages co-circulate and thus provide conditions for dual infections with different subclades. Finally, the detection of an A2/D-B3 recombinant in both Europe and the US indicates this recombinant lineage has spread beyond a single country. Recombinants are generally considered rare in EV-D68, but whether the international detection signals a potentially fit gene combination or a chance detection, it emphasizes continued importance of whole-genome surveillance.

This study collected data on existing detections of EV-D68 cases at the participating sites during 2025. As a result, there was a considerable heterogeneity in which samples were tested for EV-D68 testing, as well as the practices for which patients were tested for EV/RV. In consequence, neither denominator nor case numbers were comparable across the participating sites. Nevertheless, despite this heterogeneity there was a consistency of predominance of subclade A2/D across countries.

## Conclusion

In this multicentre European collaboration, we documented a marked shift in EV-D68 epidemiology. In 2025 adults accounted for more than 60% of detected cases and subclade A2/D predominated across Europe, representing the most pronounced shift toward adult case detection that has been observed hitherto. This pattern was consistent across the participating countries and multiple surveillance systems, despite some limited variance in sampling methodology, suggesting a genuine epidemiological change.

Wastewater surveillance supported these findings independently of clinical sampling biases, while also revealing a subclade B3 diversity that was not captured by clinical detections - underscoring the complementary value of environmental and genomic surveillance for characterizing EV-D68 circulation. In addition, the identification of interclade A2/D-B3 recombinants in both Europe and the US highlights the critical need for whole-genome sequencing alongside traditional VP1 genotyping.

Continued surveillance across all age groups is essential to determine whether the 2025 shift is sustained, and to clarify the role adults play in EV-D68 circulation. To understand the inter-generational transmission dynamics of EV-D68 will require further age-stratified surveillance and serological studies to fully resolve, and will have implications for infection control and prevention.

## Data Availability

An interactive version of the Nextstrain tree shown in Fig.3 can be viewed via the Nextstrain community feature: https://nextstrain.org/community/enterovirus-phylo/evd68-2025/vp1. Metadata associated with the samples generated in this study is downloadable from this tree.
All code used in the generation of the phylogeny, age/clade analyses, and generation of age/clade figures is available at on Github at https://github.com/enterovirus-phylo/evd68-2025.
Sequences have been deposited in GenBank under the accessions listed in the manuscript.

https://github.com/enterovirus-phylo/evd68-2025

## Acknowledgments

The authors wish to thank the European Society for Clinical Virology (ESCV) for hosting the European Non-Polio Enterovirus Network (ENPEN). The authors would also like to thank everyone who contributed to the collection, testing and reporting of clinical and virological data presented in this study. Additionally, we are grateful to scientists and public health authorities globally for sharing EV-D68 sequences and metadata openly and without delay. services.

For the Netherlands’ GP sentinel surveillance (NL1b) we thank: Lynn Aarts, Sanne Bos, Jasper van den Brink, Sharon van den Brink, Jeroen Cremer, Dirk Eggink, Zandra Felix Garza, Rianne van Gageldonk, Gabriel Goderski, Maxime Hartwig, Anne Huiberts, Jonne Mensinga-Kruize, Tara Sprong, Anne Teirlinck, Mariam Bagheri, Samantha Zoomer, Michelle van den Oever, molecular pool and virus isolation and characterisation technicians, National Institute for Public Health and the Environment (RIVM), Bilthoven; Nivel Primary Care Database – Sentinel Practices team, Mariette Hooiveld, Anneke Taal, Christy Simons, Marloes Hellwich, Cathrien Kager, Lucy Overbeek, participating general practices and their patients, Nivel, Utrecht.

For the Swedish site (SE1) thank the Clinical Genomics Stockholm facility at the Science for Life Laboratory for expertise and support with next generation sequencing

Claude Haiku 5.5 was used sparingly to aid in rephrasing some sentences for clarity and length, but not write content.

## Data Availability

An interactive version of the Nextstrain tree shown in Fig. 3 can be viewed via the Nextstrain community feature: https://nextstrain.org/community/enterovirus-phylo/evd68-2025/vp1. All code used in the generation of the phylogeny, age/clade analyses, and generation of age/clade figures is available at on Github at https://github.com/enterovirus-phylo/evd68-2025.

Sequences have been deposited in GenBank under accession numbers: PZ012370 to PZ012392, PZ012394 to PZ012396 (IS1); PZ576553 to PZ576556 (IT1 & IT4); PZ602616 (IT2); PZ621856 (IT4); PZ645974 to PZ645977 (UK2); PZ655194 (SI2); PZ672094 to PZ672107 (DK1); PZ670040, PZ670041 (FR2); PZ672960 to PZ672969 (SE1); PZ685471 to PZ685494 (ES2); PZ679146 (DE1); PZ697379 to PZ697383 (NL2); PZ946875 and PZ946876 (PT1). Sequences from sites BE1, BE2, and UK3 are not yet uploaded but will be uploaded for submission to a journal.

## Conflict of interest

None declared.

## Ethical statement

Ethical approval was obtained as required by local regulations (ES2: Hospital Universitari Vall d’Hebron Clinical Research Ethics Committee, PR(AG)419/2023; FR02: Institutional Review Board IRB00013412, number 2024-CF393; IT3: Ethics Committee of Verona and Rovigo provinces, Prot. n. 68160 (24/11/2025); SI1: Slovenian National Medical Ethics Committee, 0120-472/2024-2711-3; SI2: Slovenian National Medical Ethics Committee, 0120-384/2025-2711-3; SE1: Swedish Ethical Review Authority, 2024-02251-01). NL1b: the Centre for Clinical Expertise at the RIVM (IDS-688) declared that for (pseudo)anonymized research based on GP sentinel surveillance prior ethical approval is not required. BE2: As national reference centre for enteroviruses, the role to perform epidemiological surveillance and scientific research is described in the Royal Decree of 9/2/2011.

## Funding statement

The funders had no role in study design, data collection and analysis, decision to publish, or preparation of the manuscript. AA: Research was partially supported by Fondo de Investigación Sanitaria, Ministerio Español de Economía y Competitividad (grant FIS PI22/00023). APir: Centre for Disease Prevention and Control (CCM) of the Italian Ministry of Health (Echo11 Project, program 2023, J69I23002050001). CC, EPo, RF: EU funding within the MUR PNRR Extended Partnership Initiative on Emerging Infectious Diseases (Project No. PE00000007, INF-ACT); Italian Ministry of Health “Fondi Ricerca Corrente” L1P3. EBH: This work was supported by a Swiss National Science Foundation Starting Grant (TMSGI3_211225). Additionally, this project was supported by the ESCAPE project, funded by the European Union (101095619). Views and opinions expressed are however those of the author(s) only and do not necessarily reflect those of the European Union or European Health and Digital Executive Agency (HADEA). Neither the European Union nor the granting authority can be held responsible for them. This work has received funding from the Swiss State Secretariat for Education, Research and Innovation (SERI) (contract number 22.00482). EPa: Next Generation European Union-Ministero Universitá e Ricerca Piano Nazionale Ripresa e Resilienza (PNRR) extended partnership initiative on emerging infectious diseases (grant number PE00000007, One Health Basic and Translational Research Actions addressing Unmet Needs on Emerging Infectious Diseases INF-ACT). KBi, PCr, PM: UK Clinical Virology Network. LC, EW: The national reference centre for enteroviruses in Belgium is supported by the public health institute Sciensano (through funding of RIZIV/INAMI). NB: The work is performed as part of the National ILI and ARI surveillance and is supported by the MoH Republic of Slovenia. RD: Partially funded by Swedish Foundation for Research and Development in Medical Microbiology grant (20190318). TU: Slovenian Research and Innovation Agency (P3-0083).

## Supplement

(see files at end)

## Supplementary Methods

**Table S1.** Affiliations, surviellance type, and type of laboratory and institute for participating sites.

| Site | Affiliation(s) | Surveillance | Laboratory type | Institution type |
| --- | --- | --- | --- | --- |
| BE1 | Laboratory Medicine, Medical Microbiology, AZ Sint-Jan Brugge, Brygge, Belgium | Combination of EV clinical and ILI/ARI | Clinical | Public hospital (tertiary care) |
|  | > Department of Laboratory Medicine, National reference centre for enteroviruses, University Hospitals Leuven, Leuven, Belgium |  |  |  |
| BE2 | > Department of Microbiology, Immunology and Transplantation, Rega Institute, Laboratory of Clinical and Epidemiological Virology, KU Leuven, Leuven, Belgium | EV clinical | Clinical | Academic hospital |
| DE1 | Institute of Virology, University Hospital Bonn, Bonn, Germany | ILI/ARI | Clinical | Academic hospital |
| DK1 | Statens Serum Institut, Copenhagen, Denmark | EV clinical, ILI | Public Health | Public health |
|  | > Institut Recerca Pediàtrica, Hospital Sant Joan de Deu, Barcelona, Spain |  |  |  |
|  | > CIBER of Epidemiology and Public Health (CIBERESP), Madrid, Spain |  |  |  |
| ES1 | > Universitat Internacional de Catalunya, Barcelona, Spain | EV clinical (pediatrics) | Clinical | Academic hospital |
|  | > Respiratory Viruses Unit, Microbiology Department, Vall d'Hebron Hospital Universitari, Vall d'Hebron Institut de Recerca (VHIR), Barcelona, Spain |  |  |  |
|  | > Pediatric Hospitalization Unit, Hospital Infantil i de la Dona Vall d'Hebron, Barcelona, Spain |  |  |  |
| ES2 | > Infection and Immunity in Pediatric Patients Research Group, Vall d'Hebron Institut de Recerca (VHIR), Barcelona, Spain | EV clinical | Clinical | Academic hospital |
| FR1 | National Reference Center for Enteroviruses, Hospices Civils de Lyon, Lyon, France | EV clinical | Clinical | Academic hospital |
| FR2 | CHU Clermont-Ferrand, Centre National de Référence des entérovirus, Clermont-Ferrand, France | EV clinical | Clinical | Academic hospital |
| IS1 | Department of Clinical Microbiology, Landspítali, Reykjavik, Iceland | EV clinical | Clinical & Public Health | Academic hospital and Public health laboratory |
| IT1 | Department of biomedical sciences for health, University of Milan, Milan, Italy | ILI | Public Health | Public health |
| IT2 | Genomics and Bioinformatics Laboratory, Microbiology and Virology Department, Fondazione IRCCS Policlinico San Matteo, Pavia, Italy | EV clinical | Clinical | Hospital |
| IT3 | Department of Infectious, Tropical Diseases and Microbiology, IRCCS Sacro Cuore Don Calabria Hospital, Negrar di Valpolicella, Verona, Italy | ILI/ARI | Clinical | Hospital |
| IT4 | Clinical Microbiology and Virology Unit, ASST BRIANZA, Vimercate, Italy | ILI/ARI | Clinical | Public Hospital |
|  | > National Institute for Public Health and the Environment, Bilthoven, the Netherlands |  |  |  |
|  | > Reinier Haga Medisch Diagnostisch Centrum, Delft, the Netherlands |  |  |  |
|  | > Microvida, Laboratory for Medical Microbiology and Immunology, Tilburg, the Netherlands |  |  |  |
| NL1a | > Maasstad ziekenhuis, Rotterdam, the Netherlands | EV surveillance | Public Health | Public health (cases from hospital) |
| NL1b | National Institute for Public Health and the Environment, Bilthoven, the Netherlands | ILI/ARI, GP sentinel | Public Health | Public health |
| NL2 | Department of Viroscience, Erasmus University Medical Center, Rotterdam, the Netherlands | EV clinical | Clinical | Academic hospital |
| NO1 | Norwegian Institute of Public Health, Oslo, Norway | EV clinical and AFP | Public Health | Public health |
| PT1 | National Reference Laboratory for Influenza Virus and Other Respiratory Viruses, National Institute of Health Doutor Ricardo Jorge, Lisbon, Portugal | ILI/ARI | Public Health | Public health institute |
|  | > Department of Clinical Microbiology, Karolinska University Hospital, Stockholm, Sweden |  |  |  |
| SE1 | > Department of Microbiology, Tumor and Cell Biology (MTC), Karolinska Institutet, Stockholm, Sweden | EV clinical | Clinical | Academic hospital |
| SI1 | National Laboratory of Health, Environment and Food, Department for Public Health Microbiology, Maribor, Slovenia | ILI/ARI | Public Health | Public health |
| SI2 | Institute of Microbiology and Immunology, Faculty of Medicine, University of Ljubljana, Slovenia | EV clinical | Clinical | Academic hospital |
| UK1 | NHS Lothian, Edinburgh, United Kingdom | EV clinical | Clinical | Hospital |
|  | > Wolfson Centre for Global Virus Research, School of Life Sciences, University of Nottingham, Nottingham, United Kingdom |  |  |  |
| UK2 | > Portsmouth Hospitals University NHS Trust, Portsmouth, United Kingdom | ILI/ARI (children, immunocompromised, and ITU) | Clinical | Academic hospital |
| UK3 | WHO Global Specialised Laboratory for Polio, Medicines and Healthcare Products Regulatory Agency, South Mimms, United Kingdom | Environmental (wastewater) | Public Health | Public health research institute |

### 0.1 Phylogenetic analysis

11 sequences were excluded from phylogenetic analysis due to length *≤*300bp. All 11 sequences were able to be reliably subtyped using Nextclade, and were included in non-phylogenetic analyses (see age analysis, below).

13 short sequences had an exceptionally high number of mutations and/or ambiguous bases at the beginning and/or end of the sequence, when compared to the nearest sequence in Nextclade (e.g. 1+ mutation in every codon triplet for sequential 21 codons). This is likely due to sequencing and/or assembly issues rather than real biologic diversity, and some of the bases were flagged during Genbank submission as being 100% identical to Illumina adapter sequences. To remove this influence on the phylogeny, sequences were manually inspected, and those displaying such divergence had bases prior to VP1 nucleotide 130 and after VP1 nucleotide 490 trimmed. This led to up to 75 bases being trimmed (mean: 26 bases) from these sequences. All were then included in the phylogenetic analyses except HUJT-198343418 which was <300bp.

### 0.2 Age analysis

All sequences included in the phylogenetic analysis were included in the age analysis and figure, if age could be classified into *≥*18 and <18 years.

In addition, sequences that were <300bp but could be confidently assigned a genotype using Nextclade were included, which allowed all 11 of the 2025 sequences excluded from the phylogenetic analysis to be included. This lead to a total of 114 clinical samples from 2025 with age data being included.

### 0.3 Recombination analysis

RDP5 was used to investigate the sequences SWE/08 and NL/GP-01 for recombination. A set of 44 whole genome sequences (Table S2) were manually chosen from the curated Nextstrain dataset to represent diversity across the phylogenetic tree (Figure S1). Each of the sequences were run as a query against this reference dataset using default settings. A recombination signal was detected by seven of nine methods (Table S3). Simplot++ was then used to generate similarity plots against the parental lineages, using the configuration window: 200 base pairs, step: 30 base pairs, model: Kimura 2-parameters (Figure S2).

**Table S2.**
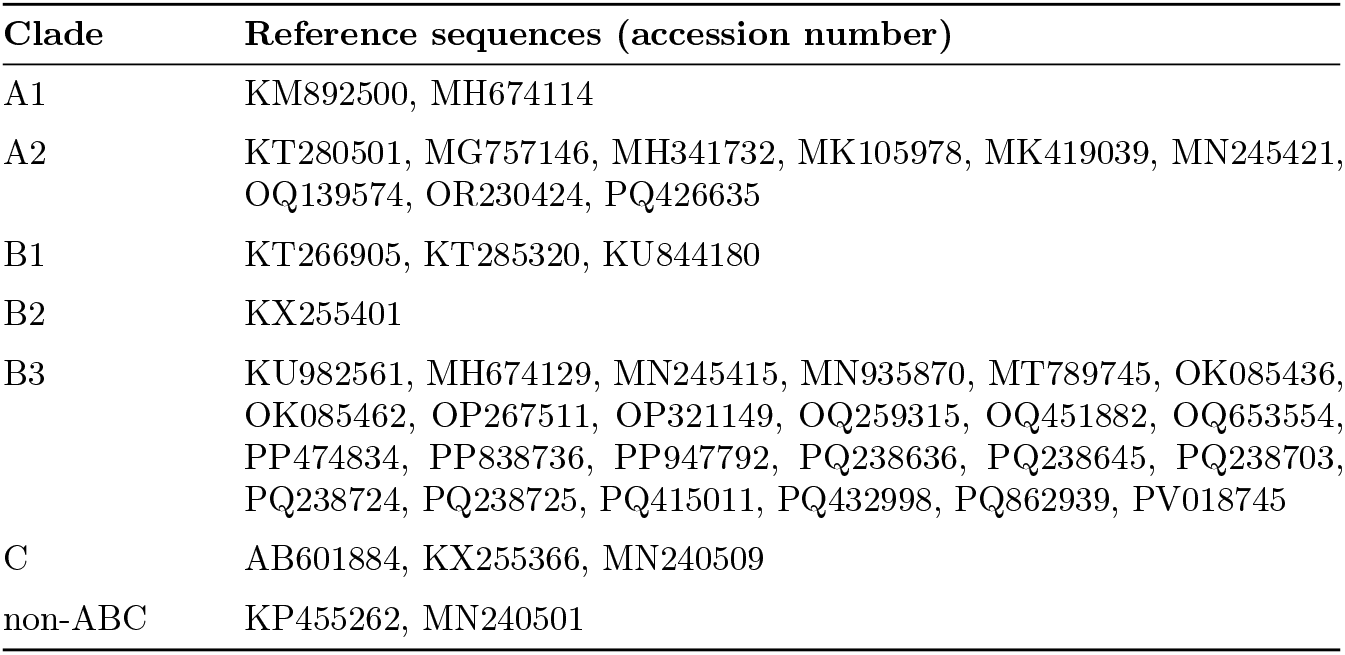
Reference sequences for recombination analysis.

| Clade | Reference sequences (accession number) |
| --- | --- |
| A1 | KM892500, MH674114 |
| A2 | KT280501, MG757146, MH341732, MK105978, MK419039, MN245421, OQ139574, OR230424, PQ426635 |
| B1 | KT266905, KT285320, KU844180 |
| B2 | KX255401 |
| B3 | KU982561, MH674129, MN245415, MN935870, MT789745, OK085436, OK085462, OP267511, OP321149, OQ259315, OQ451882, OQ653554, PP474834, PP838736, PP947792, PQ238636, PQ238645, PQ238703, PQ238724, PQ238725, PQ415011, PQ432998, PQ862939, PV018745 |
| C | AB601884, KX255366, MN240509 |
| non-ABC | KP455262, MN240501 |

**Table S3.** RDP5 results.

| Breakpoint<br>Positions In<br>Recombinant<br>Sequence |  | Recombinant Sequence | Minor Parental Sequence | Major Parental Sequence | Detection Methods |  |  |  |  |  |  |  |  |
| --- | --- | --- | --- | --- | --- | --- | --- | --- | --- | --- | --- | --- | --- |
| Begin | End |  |  |  | RDP | GENECONV | Bootscan | Maxchi | Chimaera | SiScan | PhylPro | LARD | 3Seq |
| 710* | 3728 | SWE/EVD68_08/2025 | ref_A2_PQ426635 | ref_B3_PQ415011 | 3.64E-82 | 4.50E-74 | 1.13E-79 | 1.19E-46 | 5.00E-48 | 8.94E-53 | NS | NS | 3.46E-161 |
| 3598* | 6817 | EV-D68/Netherlands/<br>GP01/2025 | ref_B3_OP267511 | ref_A2_PQ426635 | 4.95E-55 | 5.55E-53 | 7.20E-42 | 9.65E-40 | 1.59E-38 | 2.49E-45 | NS | NS | 3.99E-99 |

\* = The actual breakpoint position is undetermined

**Fig S1.**
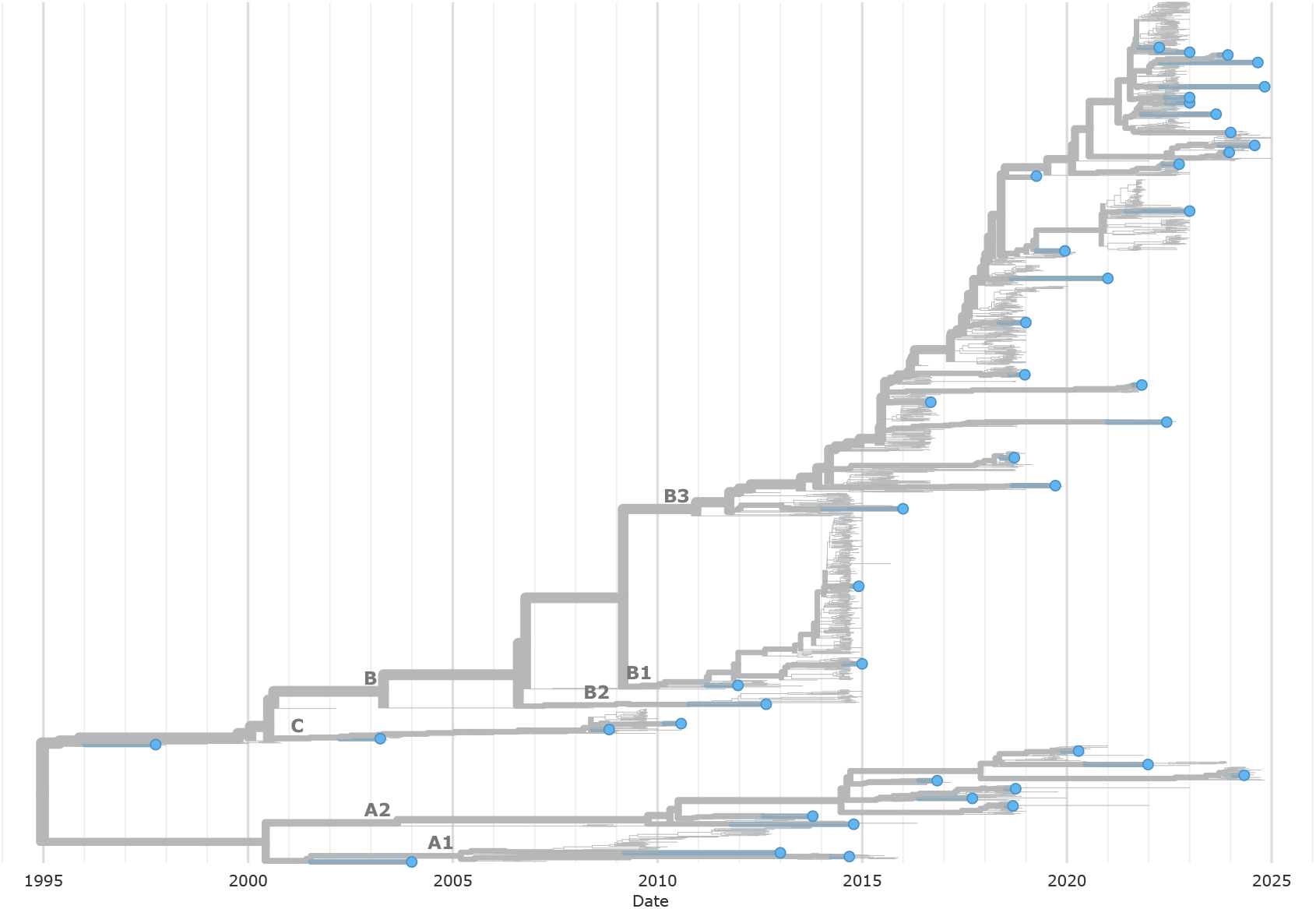
Reference set for recombination analysis. For the recombination analysis, a reference set of 44 whole genome sequences was manually selected from different parts of the phyologenetic tree, with the aim to ensure representativeness. Blue dots represent the chosen reference sequences.

**Fig S2.**
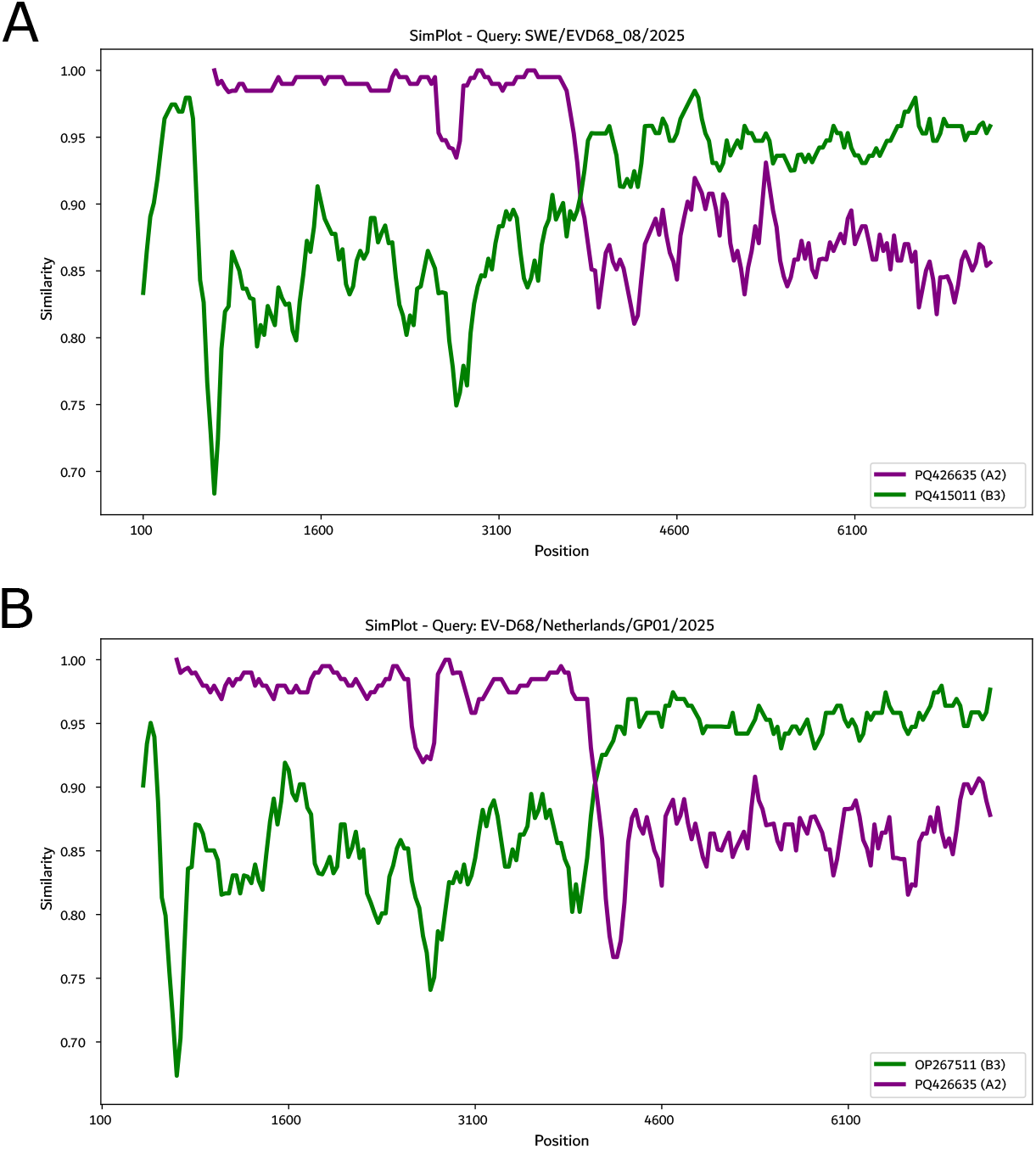
Similarity plot. Similarity plots against A2 (purple) and B3 (green) sequences. A: The recombinant sequence from Sweden has the 5’-part derived from subclade A2/D and 3’-part from subclade B3, with a tentative breakpoint at approximately 3,750 bp. B: The recombinant sequence from the Netherlands also has the 5’-part derived from subclade A2/D and 3’-part from subclade B3, with a tentative breakpoint at approximately 3,600 bp. Plots generated using Simplot++.

**Fig S3.**
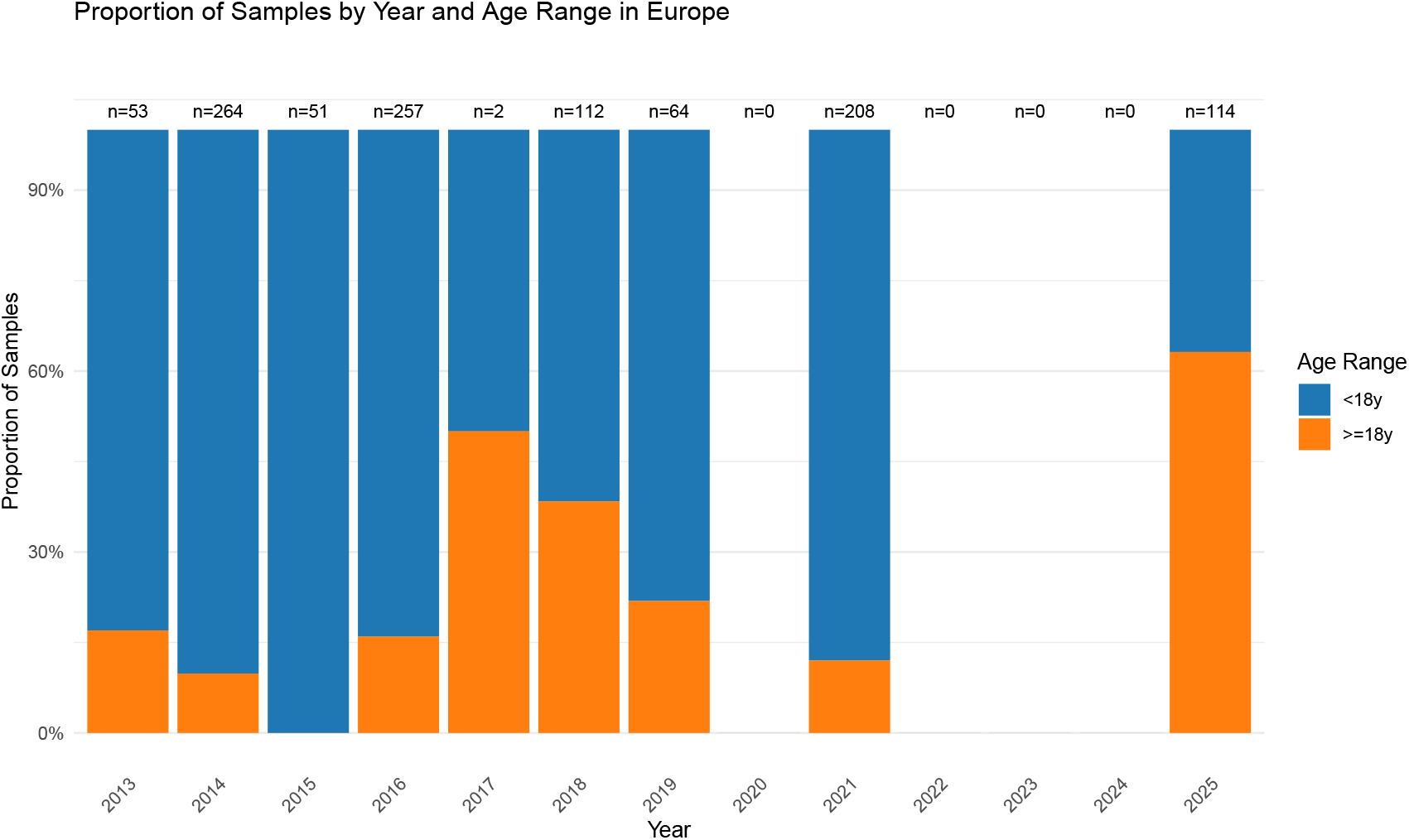
Proportion of sequences from children vs adults 2014-2025. European sequences with at least 300bp of VP1, grouped by age (where available) and year. In non-outbreak years, number of samples can be very small and should be interpreted with caution. In some years, no age data is available without manual or semi-manual curation, which has not been performed (this data used to be curated by ViPR which is now defunct); age data is available for *≥*100 samples for all years compared in the main text (2016, 2018, 2021, 2025).

**Fig S4.**
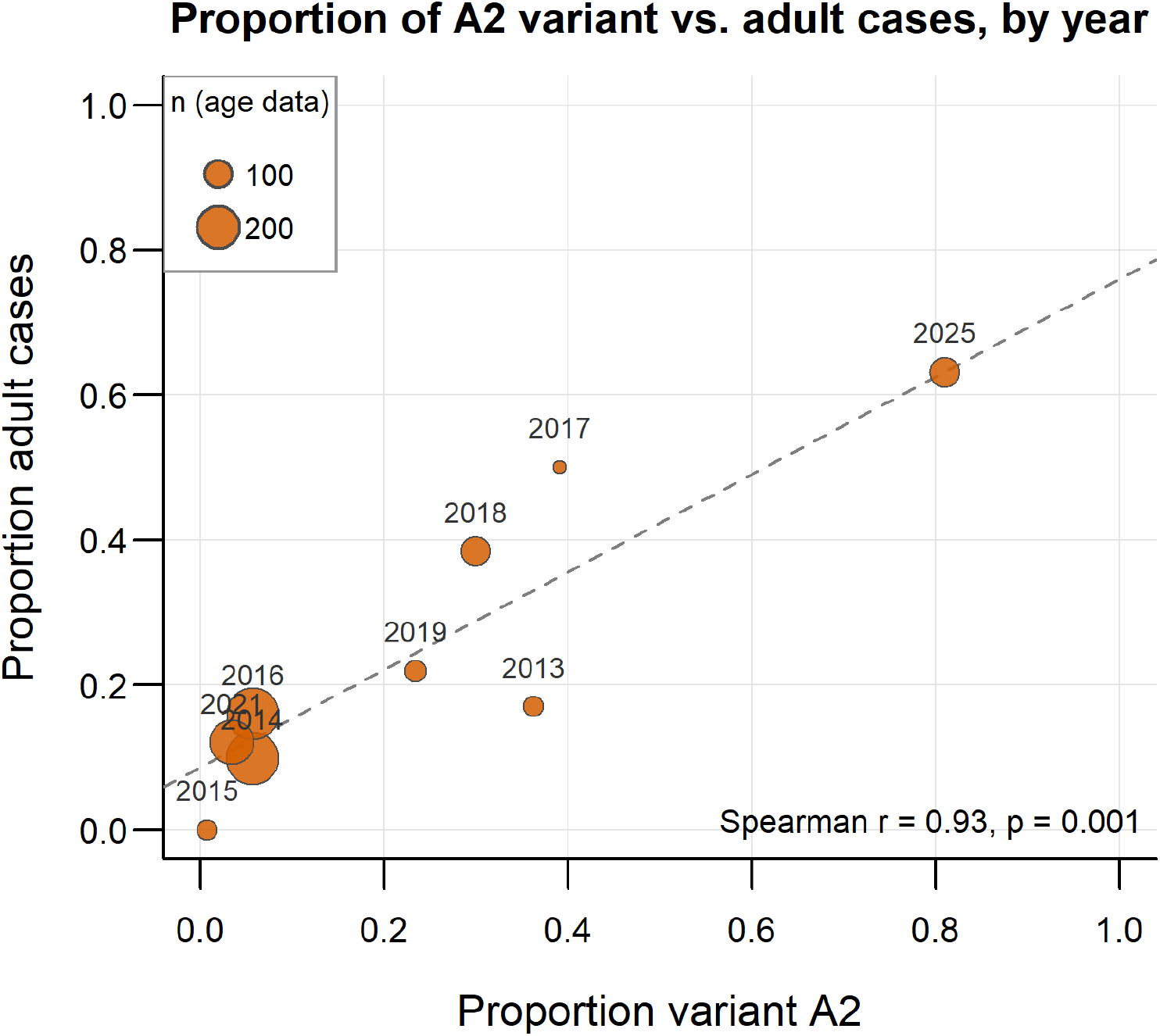
Proportion of sequences with A2 subgenogroup vs. adult cases. For years since 2013, the proportion of sequences with available age is plotted against the proportion of adult ( *≥*18 years-old) cases. Though numbers are small for some years and should be interpreted with caution, there is a general trend towards years with a higher A2 proportion having more adult cases.

## Notes

### Competing Interest Statement

The authors have declared no competing interest.

### Author Declarations

Hospital Universitari Vall d'Hebron Clinical Research Ethics Committee gave ethical approval for this work (PR(AG)419/2023). CHU Clermont-Ferrand Centre National de Reference des enterovirus Institutional Review Board IRB00013412 gave ethical approval for this work (2024-CF393). Ethics Committee of Verona and Rovigo province gave ethical approval for this work (Prot. n. 68160 (24/11/2025)). Slovenian National Medical Ethics Committee gave ethical approval for this work (0120-472/2024-2711-3 and 0120-384/2025-2711-3). Swedish Ethical Review Authority gave ethical approval for this work (2024-02251-01). The Centre for Clinical Expertise at the RIVM waived ethical approval for this work (as (pseudo)anonymized research based on GP sentinel surveillance does not require ethical approval). The Department of Laboratory Medicine at University Hospitals Leuven and Laboratory of Clinical and Epidemiology Virology at KU Leuven waived ethical approval for this work (as national reference centre for enteroviruses, the role to perform epidemiological surveillance and scientific research is described in the Royal Decree of 9/2/2011).

